# The quality and reliability of short videos about External Counterpulsation on TikTok: a cross-sectional study

**DOI:** 10.64898/2026.02.22.26346843

**Authors:** Shengkun Gai, Dongqi Li, Glen Borchert, Fangwan Huang, Xiuyu Leng, Jingshan Huang

**Affiliations:** Department of Cardiology, Linfen People’s Hospital, Linfen, Shanxi, China; Department of Preventive Medicine, Feinberg School of Medicine, Northwestern University, Chicago, IL, U.S.A; Department of Pharmacology, College of Medicine, University of South Alabama, Mobile, AL, U.S.A; College of Computer and Data Science, Fuzhou University, Fuzhou, China; Department of Cardiology, The First Affiliated Hospital, Sun Yat-sen University, Guangzhou, China; NHC Key Laboratory of Assisted Circulation and Vascular Diseases (Sun Yat-sen University), Guangzhou, China; Department of Computer Science, School of Computing, University of South Alabama, Mobile, AL, U.S.A

**Keywords:** EECP, TikTok, Social media, Health information quality, Patient education

## Abstract

**Background:** Short-video platforms have become increasingly important sources of health information for the general public. However, the informational quality and dissemination patterns of content related to specific therapeutic modalities, such as enhanced external counterpulsation (EECP), remain insufficiently characterized. This study aimed to evaluate the informational quality of EECP-related videos on a short-video platform and to examine the relationship between content quality and user engagement.

**Methods:** A cross-sectional content analysis was conducted on EECP-related short videos identified through keyword-based searches. Informational quality was independently assessed using four validated instruments: the Global Quality Scale (GQS), the Journal of the American Medical Association (JAMA) benchmark criteria, the modified DISCERN instrument (mDISCERN), and the Video Information and Quality Index (VIQI). Video characteristics and user engagement metrics were extracted and analyzed.

**Results:** Overall, EECP-related videos demonstrated low-to-moderate informational quality across all assessment tools. Longer video duration was consistently associated with higher informational quality scores. In contrast, user engagement metrics, including the number of likes and comments, showed weak or negative associations with informational quality. Compared with videos addressing other coronary heart disease treatments, EECP-related videos were less frequently represented and received lower overall engagement.

**Conclusions:** EECP-related content on short-video platforms is characterized by limited visibility and modest informational quality, with a notable misalignment between user engagement and informational value. These findings suggest that clinically relevant but complex therapies such as EECP may be structurally disadvantaged in short-video health communication environments.

## Background

Coronary heart disease remains one of the leading causes of morbidity and mortality worldwide, imposing a substantial burden on patients and healthcare systems [1,2]. In addition to pharmacological treatment and revascularization strategies, non-invasive therapeutic approaches have been developed to improve symptoms and quality of life in selected patient populations. Enhanced external counterpulsation (EECP) is a non-invasive therapeutic technique that applies sequential external pressure to the lower extremities during diastole, with the aim of enhancing diastolic coronary perfusion and reducing cardiac workload [3].

EECP has been reported to be associated with improvements in clinical symptoms, exercise tolerance, and quality of life among patients with refractory angina and other forms of coronary heart disease. Its therapeutic mechanisms are thought to involve hemodynamic effects, endothelial function improvement, and the promotion of collateral circulation. Owing to these characteristics, EECP has been incorporated into clinical practice and guideline recommendations in certain settings, particularly for patients who are not suitable candidates for invasive interventions [3].

Prior literature provides both clinical and communication-context foundations for the present analysis. As examples of clinical foundations, an expert consensus summarizing indications, contraindications, and practical considerations for EECP in elderly populations was published in 2019 [4] and a long-term follow-up study examining outcomes of EECP treatment in 1,427 patients suffering from chronic refractory angina and/or related coronary syndromes was published in 2008 [5]. As examples of communication-context foundations, the quality and educational value of oral-health content on short-video platforms has been evaluated using established assessment approaches [6]. In addition, short-video platforms such as TikTok have been examined as channels for health-related messaging, including COVID-19 communication, and public engagement [7].

Of note, classic criteria and benchmarks have been proposed to assess the quality of online medical information [8]. Subsequent work has applied structured benchmarks to video-based patient education materials to evaluate reliability and completeness [9]. Across multiple clinical topics, video-content analyses on platforms such as YouTube have repeatedly found gaps between popularity and informational quality [10,11]. In China, evaluations of hypertension/diabetes information on WeChat and TikTok (Douyin) and analyses of oral diabetes medication videos on Douyin similarly indicate heterogeneity in quality [12,13]. Short-video apps are increasingly used as health information sources for chronic diseases, raising questions about information visibility, engagement, and equity of exposure [14,15]. Case studies suggest TikTok can improve accessibility of science communication, but content framing and evidence support remain critical [16]. While health-related content on TikTok can be highly engaging, evidence support across conditions such as ADHD ranges from valid to wholly inaccurate [17]. Finally, while health misinformation on social media has been identified as a public-health challenge [18], short video platforms can unquestionably influence patients’ healthcare decisions such as selecting a physician or even when to seek care, underscoring the role of perceived credibility in digital health environments [19].

Clinical guidelines provide the broader context for employing EECP as an adjunctive option for symptomatic management in patients with stable coronary artery disease [20]. Further, guidelines outlining diagnostic and management pathways for stable ischemic heart disease, within which adjunctive non-invasive options like EECP may be considered are also available [21]. In addition, a number of expert consensus statements further refining clinical recommendations [4] and reviews summarizing patient selection, clinical uses of EECP and the use of EECP in emerging applications (e.g., EECP use in unrevascularizable patients) have also been reported [22–24].

With the rapid growth of digital media, short-video platforms have emerged as prominent channels for health information dissemination [25]. These platforms offer high accessibility and broad reach, enabling users to obtain medical information through brief, visually engaging content [26]. Consequently, short videos have become an influential medium shaping public perceptions of diseases and treatment options [6–7].

However, the structural characteristics of short-video platforms may also limit the effective communication of complex medical information. Time constraints, algorithm-driven content prioritization, and emphasis on viewer engagement can favor simplified narratives over comprehensive or evidence-based explanations. For medical interventions such as EECP, which require contextual understanding of indications, mechanisms, and limitations, these constraints may affect both the quality and visibility of information presented to the public [15].

Against this background, short-video platforms have become an increasingly important channel for disseminating health-related information to the public. However, the characteristics of short-video formats—such as limited duration, algorithm-driven content distribution, and emphasis on user engagement—may pose challenges for the communication of complex or non-mainstream medical interventions. EECP, as a non-invasive therapy with specific indications and mechanisms, represents a particularly relevant case for examining these challenges [16].

Despite its established clinical application, the extent to which EECP-related information is adequately represented and communicated on short-video platforms remains unclear [27]. Existing studies have primarily focused on the general quality of online health information, while evidence regarding the visibility, informational quality, and comparative representation of EECP in short-video environments is limited [28–31].

Therefore, this study aimed to systematically evaluate the information quality, content characteristics, and user engagement metrics of short videos related to EECP on a major short-video platform. Using validated assessment instruments, we sought to provide an objective overview of the current quality of EECP-related video content and to explore the associations between video characteristics, engagement indicators, and information quality.

## Materials and Methods

### Study Design and Video Selection

This study employed a cross-sectional content analysis design to evaluate short videos related to EECP published on a major short-video platform. Videos were identified through keyword-based searches using terms related to EECP. Only videos that were directly relevant to EECP and accessible at the time of data collection were included. Duplicate videos, non-informative clips, and content unrelated to EECP were excluded from the analysis.

In addition to topical relevance, we required that each included video contain substantive EECP-related information in spoken narration, on-screen text, or captions sufficient to permit structured quality scoring using the predefined instruments. Videos were excluded if they were duplicates, inaccessible at the time of data collection, unrelated to EECP, or lacked interpretable informational content (e.g., purely decorative clips without clinically meaningful statements). The final analytic sample consisted of 141 EECP-related short videos. For comparison, 142 videos addressing other CAD treatments were also included in the analysis.

### Data Extraction and Video Characteristics

For each included video, basic characteristics were extracted, including video duration and uploader source category. Videos were categorized based on uploader information, declared professional background, and content presented in video titles, descriptions, or captions. Uploader categories included cardiology specialists (physicians with recognized training or certification in cardiology or cardiovascular medicine), non-cardiology medical professionals (physicians from other clinical specialties), nursing professionals (registered nurses or individuals with relevant nursing qualifications), and general users without an explicitly stated medical or healthcare background. Commercial promotional accounts were classified as a separate category.

Uploader classification was determined from publicly available account information and video-level cues, including profile descriptions, verification status (when available), and self-declared professional identity presented in the video title, on-screen text, or captions. Accounts were categorized as cardiology professionals, non-cardiology medical professionals, nursing professionals, general users (no explicitly stated healthcare background), or commercial/promotional accounts. Because these indicators were self-reported and not independently verified, uploader classification may be subject to misclassification.

User engagement metrics, including the number of likes and comments, were also recorded. These variables were collected to support subsequent analyses examining the associations between video characteristics, informational quality, and audience engagement, as reported in the Results section.

### Quality Assessment and Inter-rater Reliability

The informational quality of the included videos was independently assessed using four validated instruments: the Global Quality Score (GQS), the Journal of the American Medical Association (JAMA) benchmark criteria, the modified DISCERN (mDISCERN), and the Video Information and Quality Index (VIQI). The GQS was used to assess the overall quality and usefulness of video content from a viewer’s perspective [9]. The JAMA benchmark criteria evaluated authorship, attribution, disclosure, and currency of information [8]. The mDISCERN instrument was applied to examine the completeness and reliability of treatment-related information, including discussion of benefits, risks, and supporting evidence [10]. The VIQI assessed video-specific characteristics, including flow of information, accuracy, and presentation quality [11].

All included videos were independently evaluated by two medically trained reviewers using four validated instruments: the Global Quality Score (GQS), the Journal of the American Medical Association (JAMA) benchmark criteria, the modified DISCERN (mDISCERN), and the Video Information and Quality Index (VIQI). GQS was scored on a 5-point scale from poor to excellent overall educational quality; mDISCERN consisted of five binary (yes/no) reliability items (1 point for “yes”, 0 for “no”); JAMA evaluated four benchmark domains (authorship, attribution, currency, and disclosure; 1 point per domain, total 0–4); and VIQI assessed four domains (information flow, information accuracy, video quality, and title–content precision; each 1–5, total 4–20). Detailed scoring criteria for each instrument are provided in Supplementary Tables S3–S6.

Prior to formal scoring, the two reviewers conducted a calibration exercise on a subset of videos to harmonize interpretation of the scoring criteria. After independent scoring, discrepancies were resolved through discussion to reach consensus. Inter-rater reliability for the pre-consensus continuous scores was quantified using the intraclass correlation coefficient (ICC), with higher ICC values indicating better agreement between reviewers.

### Statistical Analysis

Descriptive statistics were used to summarize video characteristics, quality scores, and user engagement metrics. Continuous variables were reported as mean ± standard deviation or median with interquartile range (IQR), as appropriate based on data distribution. Categorical variables were presented as frequencies and percentages.

Comparisons of informational quality scores across uploader categories were performed using nonparametric tests. The Mann–Whitney U test was applied for two-group comparisons, and the Kruskal–Wallis test was used for comparisons involving more than two groups. Associations between video characteristics, quality scores, and user engagement metrics were examined using Spearman’s rank correlation analysis.

All statistical analyses were performed using IBM SPSS Statistics (version 25.0). Descriptive statistics included minimum and maximum values, means with standard deviations, medians, and frequencies, as appropriate. To quantify the magnitude and direction of the effects of video parameters on assessment-tool scores, linear regression models were constructed. Correlations between variables were evaluated using Spearman’s rank correlation coefficients and were cross-validated using Kendall’s tau-b correlation coefficients to improve the robustness of the results. All statistical tests were two-sided, and a p value < 0.05 was considered statistically significant.

For transparency and reproducibility, direct links to all analyzed videos, along with their access dates, are provided in Supplementary Tables S1 (EECP group) and S2 (other CAD treatment group).

## Results

### Characteristics of External Counterpulsation Videos

A total of 141 EECP-related short videos that met the inclusion criteria were included in the analysis (Fig. 1). Descriptive statistics of video characteristics and user engagement metrics are summarized in Table 1. The duration of the included videos varied widely, ranging from 3 to 377 seconds, with a mean duration of 79.9 ± 56.4 seconds and a median of 68 seconds.

**Figure 1.**
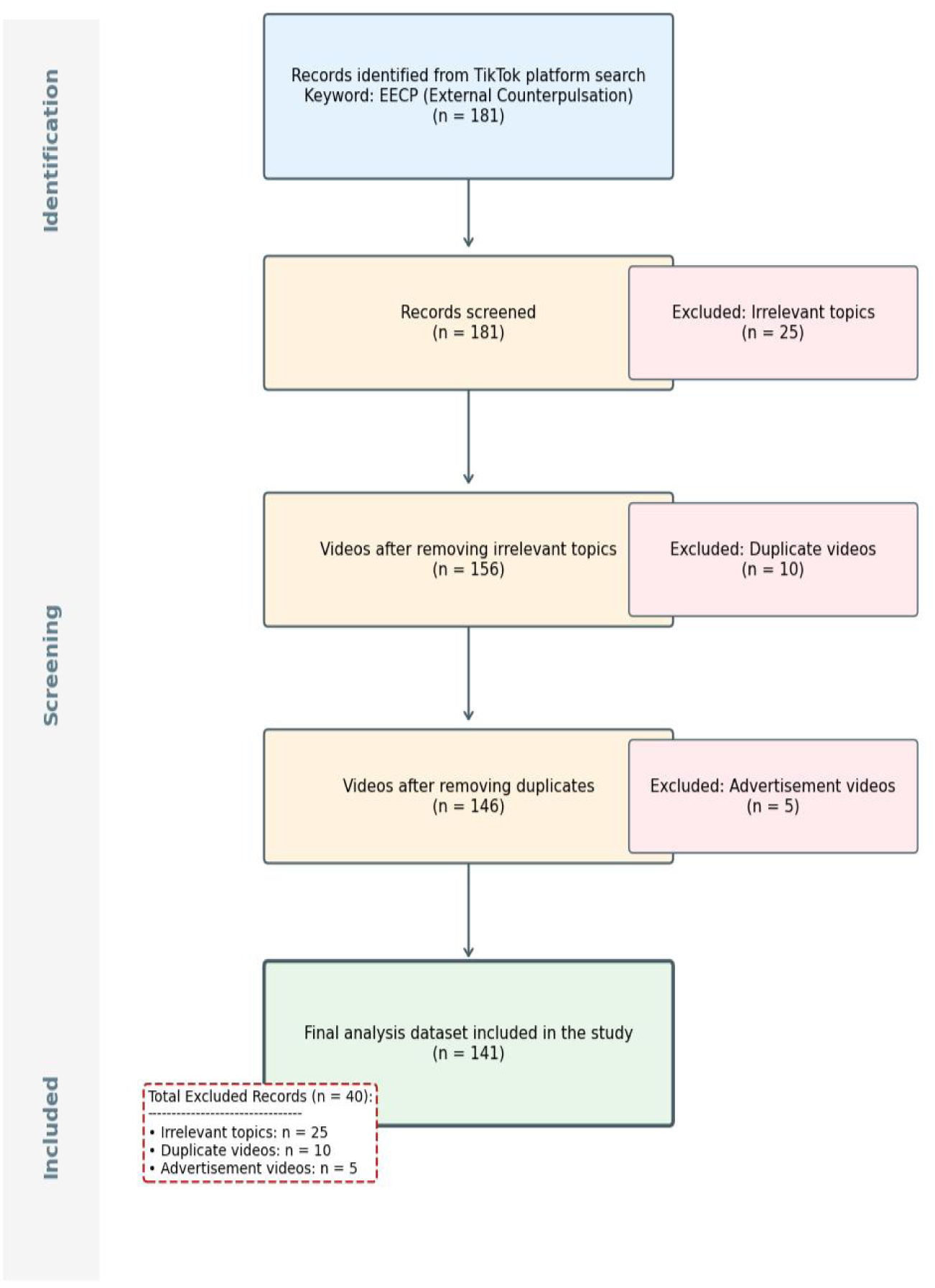
Flow chart of video selection.

**Table 1.** Descriptive statistics based on 141 short videos about EECP.

| Video parameter | Min | Max | Sum | Mean | SD | median | Quartile |
| --- | --- | --- | --- | --- | --- | --- | --- |
| Video length (s) | 3 | 377 | 11268 | 79.9149 | 56.4251 | 68 | 61.5 |
| Likes | 1 | 3678 | 24000 | 170.2128 | 499.7203 | 41 | 84 |
| Collections | 0 | 2239 | 6690 | 47.4468 | 201.4244 | 7 | 18 |
| Comments | 0 | 416 | 1886 | 13.3759 | 44.1615 | 2 | 9 |
| Shares | 0 | 1182 | 8506 | 60.3262 | 167.8814 | 9 | 30.5 |

User engagement metrics also showed substantial variability. The number of likes ranged from 1 to 3,678, with a mean of 170.2 ± 499.7 and a median of 41. The number of collections ranged from 0 to 2,239 (mean: 47.4 ± 201.4; median: 7). Comments ranged from 0 to 416, with a mean of 13.4 ± 44.2 and a median of 2, while the number of shares ranged from 0 to 1,182, with a mean of 60.3 ± 167.9 and a median of 9.

Overall, the distributions of engagement indicators were highly right-skewed, with median values substantially lower than mean values across all engagement metrics, indicating that a small proportion of videos accounted for disproportionately high levels of user interaction.

The descriptive characteristics of the included EECP-related videos are summarized in Table 1.

The distributions of video parameters and engagement metrics are shown in Fig. 2.

**Figure 2.**
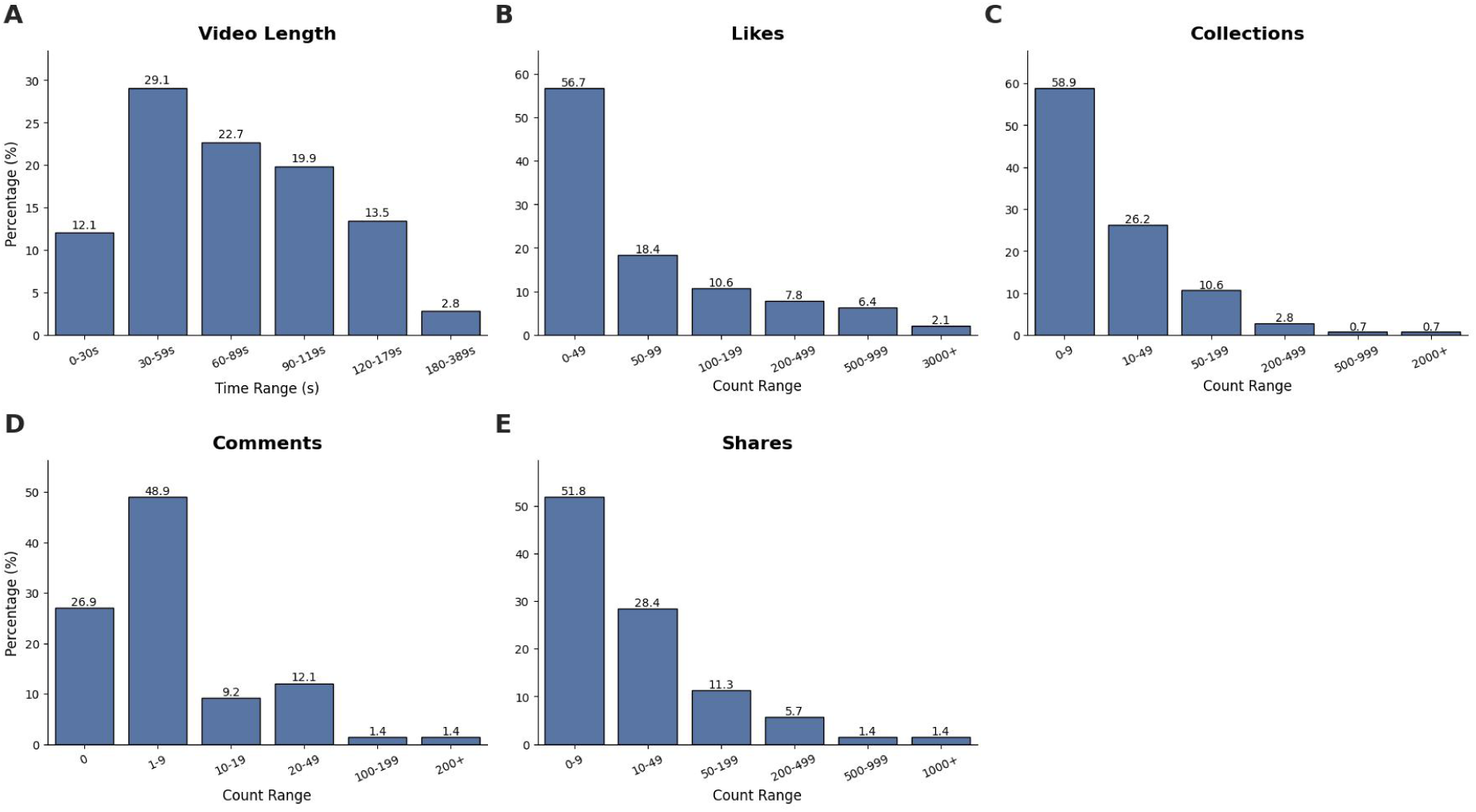
Distribution of video parameters and engagement metrics. (A) Video length (seconds). (B) Number of Likes. (C) Number of Collections. (D) Number of Comments. (E) Number of Shares. Data are presented as the percentage of total videos (N = 141). The x-axis represents the range of values for each category, and the y-axis represents the percentage of videos falling within that range. Video length shows a relatively balanced distribution, whereas engagement metrics (Likes, Collections, Comments, Shares) exhibit a right-skewed distribution, with the majority of videos receiving lower engagement counts.

### Video Quality Analysis and Evaluation

Using four validated instruments, the informational quality of EECP-related videos was systematically evaluated. Overall, scores across the Global Quality Scale (GQS), JAMA benchmark criteria, modified DISCERN (mDISCERN), and the Video Information and Quality Index (VIQI) consistently indicated low-to-moderate quality. For all four assessment tools, higher numerical scores corresponded to superior quality of the evaluated content; the score ranges were 2–4 for GQS, 1–4 for both mDISCERN and JAMA, and 5–11 for VIQI. Few videos achieved high scores simultaneously across multiple instruments, suggesting that comprehensive, balanced, and evidence-based EECP information was uncommon on the platform.

Assessment using the GQS revealed that most videos provided only limited educational value, with insufficient discussion of therapeutic indications, mechanisms, and clinical outcomes. Similarly, JAMA benchmark criteria scores demonstrated that authorship transparency, attribution of information sources, and disclosure of conflicts of interest were frequently absent. These deficiencies reduced the overall credibility of the content from a clinical perspective.

The mDISCERN assessment further showed that key elements related to treatment rationale, benefits, and potential risks were often incompletely addressed. In particular, explicit discussion of patient selection criteria and evidence supporting EECP efficacy was rarely presented. VIQI scores reflected comparable trends, indicating that while some videos were visually engaging, their informational accuracy and depth remained limited.

Taken together, the results across all four assessment tools demonstrate a consistent pattern of modest informational quality among EECP-related short videos. This convergence across independent instruments strengthens the robustness of the findings and indicates that the observed limitations are not tool-specific but reflect a broader issue in EECP-related health communication on short-video platforms.

The descriptive statistics of quality assessment scores are summarized in Table 2.

**Table 2:**
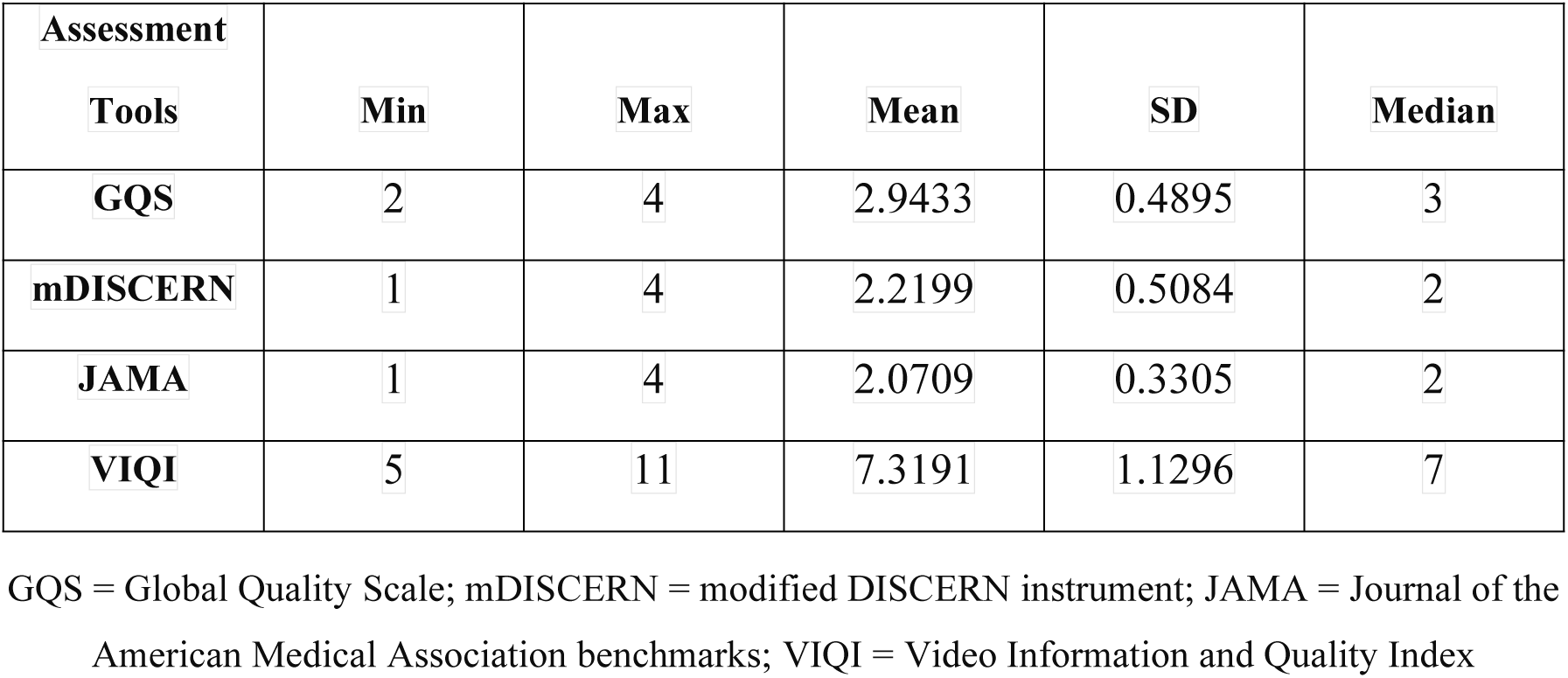
Descriptive Statistics of Quality Assessment Scores (n = 141).

The distributions of quality assessment scores are presented in Fig. 3.

**Figure 3.**
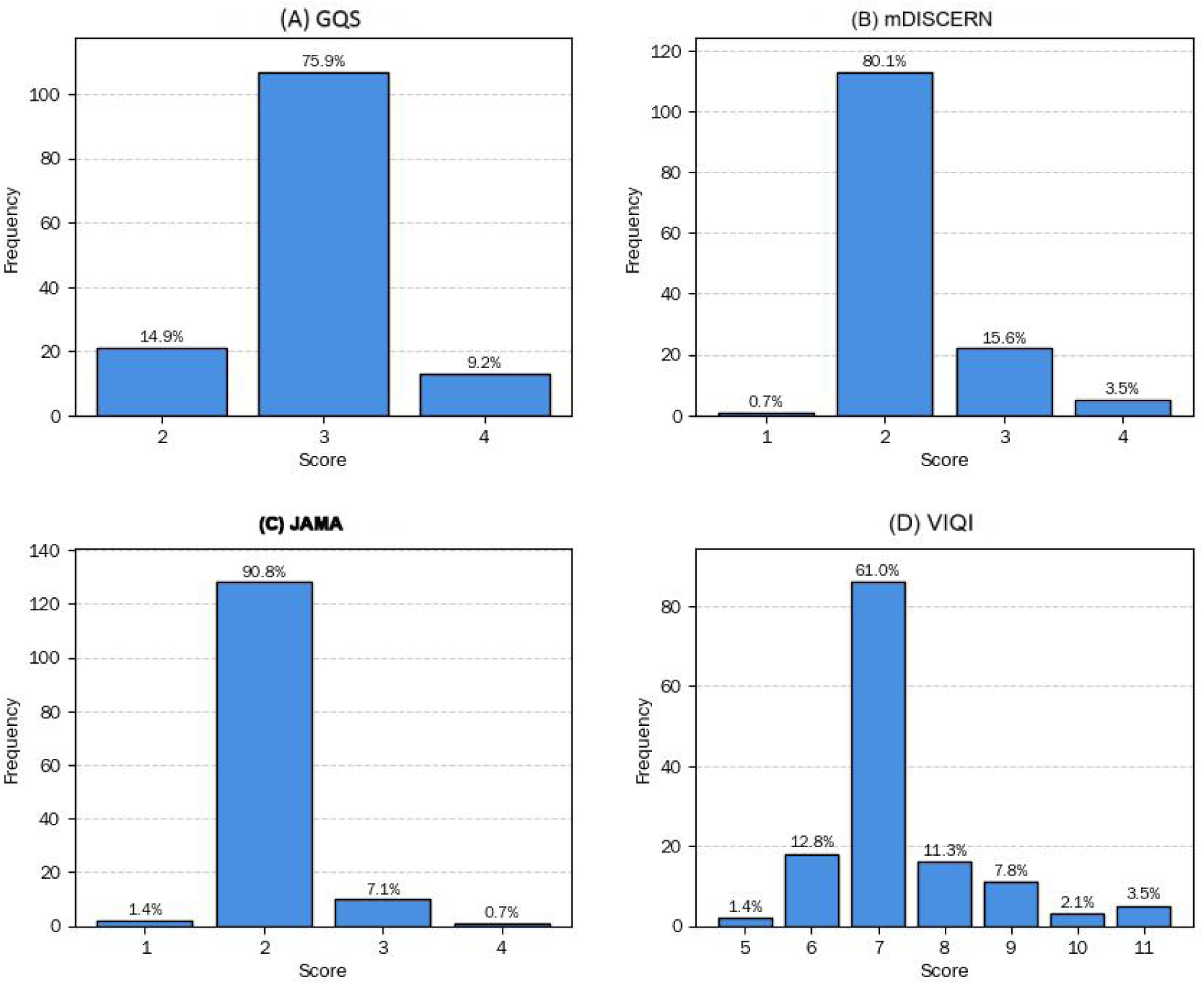
Distributions of quality assessment scores for TikTok videos related to EECP. (A) Global Quality Scale (GQS), (B) modified DISCERN, (C) JAMA benchmark criteria, and (D) Video Information and Quality Index (VIQI). Percentages shown above bars indicate relative frequencies within the dataset. Higher scores denote better video quality and reliability.

### The Influence of Video Parameters on the Score of Assessment Tools

The associations between video parameters and informational quality scores were examined across all four assessment instruments. Among the evaluated parameters, video duration demonstrated the most consistent and robust association with informational quality across all tools. Specifically, longer video duration was significantly associated with higher scores on the Global Quality Scale (GQS), JAMA benchmark criteria, modified DISCERN (mDISCERN), and the Video Information and Quality Index (VIQI) (p < 0.0001). These findings indicate that increased video length provides greater opportunity to introduce background context, explain therapeutic mechanisms, and address clinical considerations related to EECP.

In contrast, user engagement metrics showed limited or inconsistent associations with informational quality. The number of likes and comments was not positively correlated with higher quality scores and, in some analyses, exhibited weak or negative associations. These results suggest that engagement-driven popularity on short-video platforms does not reliably reflect the accuracy or completeness of medical information.

Other video parameters, such as posting time and presentation style, showed variable associations with quality scores depending on the assessment instrument used. However, these associations were generally weaker and less consistent than those observed for video duration. Collectively, the results indicate that structural characteristics of short videos—particularly constraints on video length—play a more influential role in shaping informational quality than audience-driven engagement indicators. The associations between video parameters and quality assessment scores are summarized in Fig. 4.

**Figure 4.**
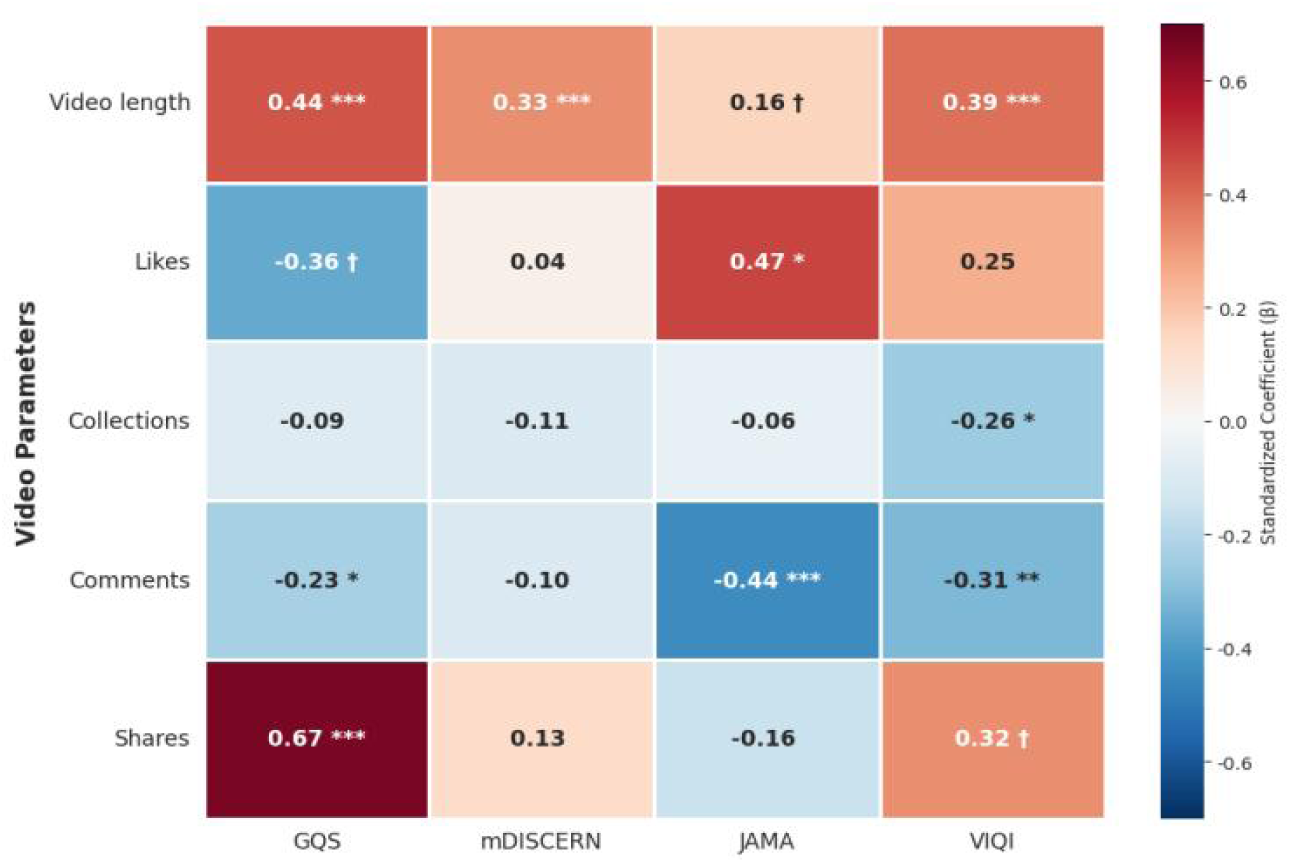
Heatmap of associations between video parameters and quality assessment scores. The color intensity represents the standardized coefficient (β), with red indicating a positive correlation and blue indicating a negative correlation. Statistical significance is denoted by symbols: ***p<0.001, **p<0.01, *p<0.05, and ^↑^p<0.1.

### Video source analysis

Videos were further analyzed according to their source to explore differences in informational quality and user engagement. Based on declared professional background inferred from account profiles, verification status, and content presented in video descriptions and captions, the included videos were categorized into the following source groups: cardiology professionals, non-cardiology medical professionals, nursing professionals, patients or general users without a stated medical or nursing background, and commercial or promotional accounts. Notable differences in informational quality and engagement metrics were observed across these source categories.

Classification of uploader professional background was based on self-reported information and publicly available account credentials (e.g., profile descriptions and verification status), rather than independent verification, and therefore may be subject to misclassification.

Videos produced by healthcare professionals generally achieved higher informational quality scores across the Global Quality Scale (GQS), JAMA benchmark criteria, modified DISCERN (mDISCERN), and the Video Information and Quality Index (VIQI) compared with videos from non-professional sources. In particular, videos uploaded by cardiology professionals more frequently provided structured explanations of EECP, addressed clinical indications and therapeutic mechanisms, and referenced evidence-based information. However, despite higher quality scores, healthcare professional–generated videos did not consistently receive higher levels of user engagement.

In contrast, videos created by patients or general users, as well as commercial or promotional accounts, tended to receive lower scores on objective quality assessments. These videos often focused on personal experiences, testimonials, or marketing-oriented narratives, with limited discussion of clinical evidence or potential limitations of EECP. Nevertheless, such videos frequently attracted greater numbers of likes and comments, indicating higher audience engagement.

The observed divergence between informational quality and user engagement across video sources underscores the influence of content origin on both educational value and audience response. While professional sources contributed higher-quality information, non-professional and promotional sources appeared to be more effective at capturing user attention on short-video platforms. The distribution of videos by uploader professional background is shown in Fig. 5.

**Figure 5.**
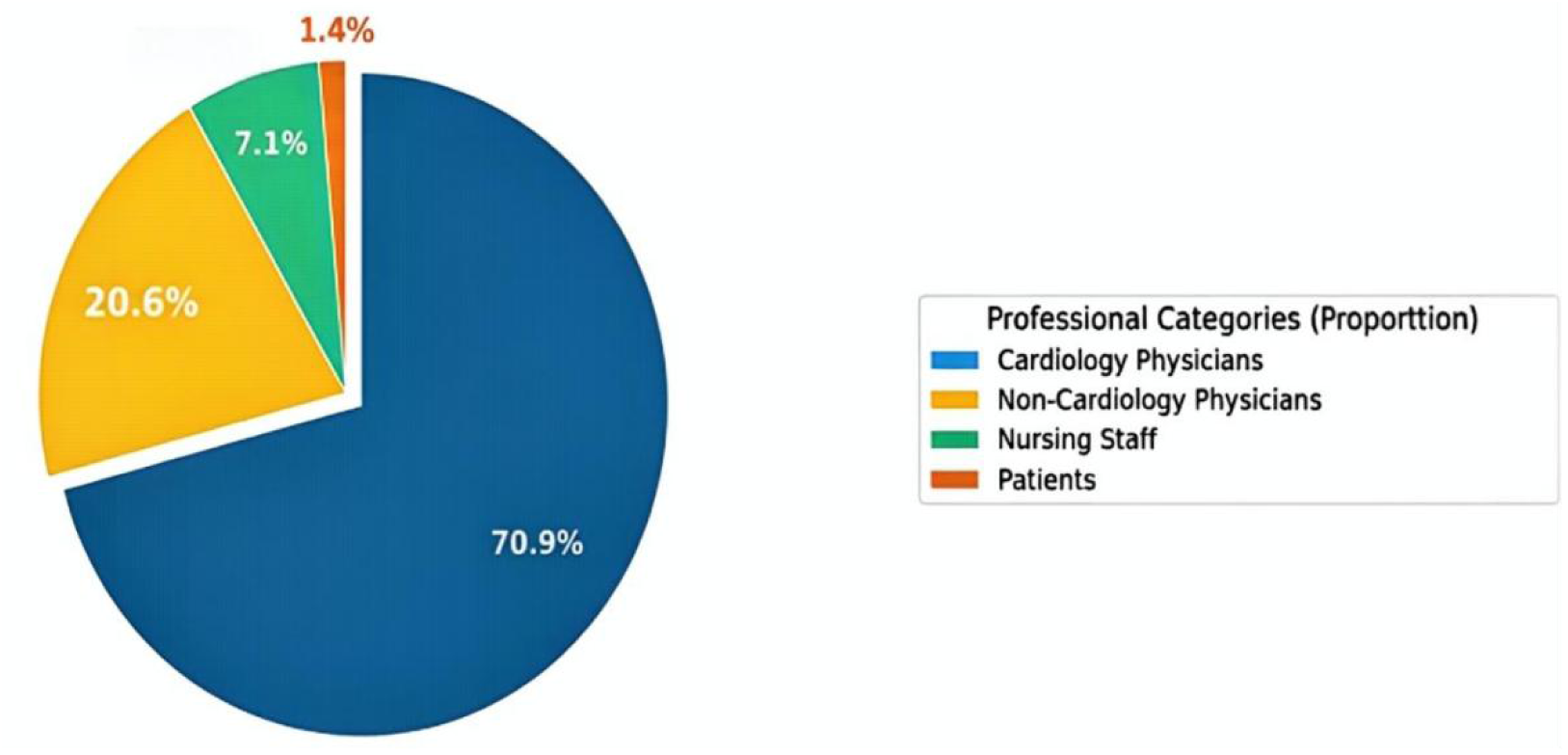
Distribution of uploader categories. The largest group consisted of cardiologists (70.9%, n=100), followed by non-cardiologist physicians (20.6%, n=29), nursing staff (7.1%, n=10), and patients (1.4%, n = 2).

A comparison of user interaction metrics between videos uploaded by cardiology and non-cardiology physicians is presented in Fig. 6. Quality assessment scores for videos uploaded by cardiology physicians and non-cardiology physicians are summarized in Table 3 and Table 4, respectively.

**Figure 6.**
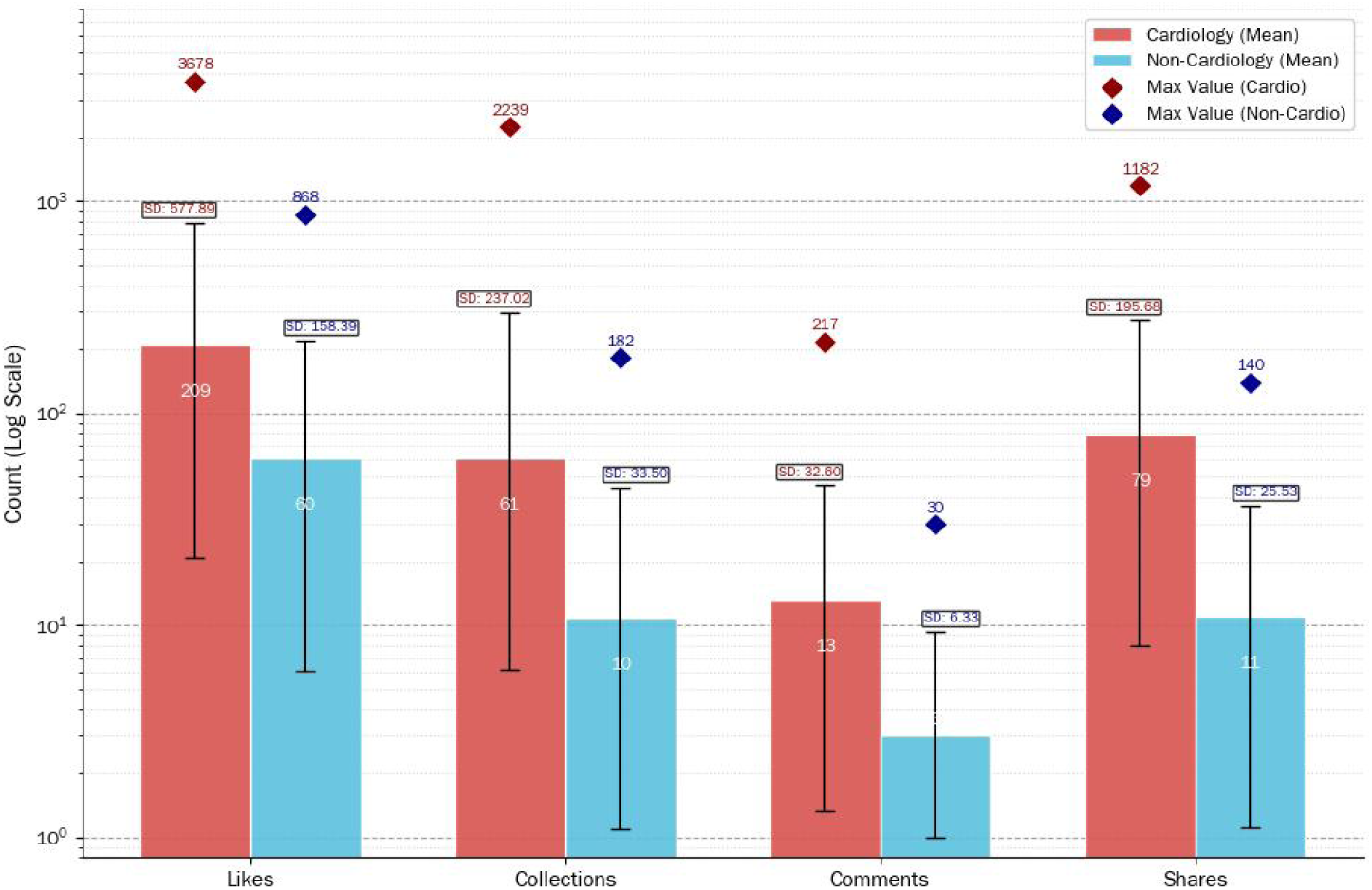
Comprehensive comparison of interaction metrics between Cardiology and Non-Cardiology videos. Bar heights represent mean values, black error bars indicate the standard deviation and diamond markers denoting the Maximum values observed. The Y-axis is presented on a logarithmic scale to accommodate the wide range of data. Cardiology videos (red) show consistently higher means, maxima, and variability across all metrics compared to Non-Cardiology videos (blue).

**Table 3.** The evaluator used four assessment tools to evaluate the scores of the videos uploaded by cardiology physicians.

| Assessment Tool | Min | Max | Median | Mean | SD |
| --- | --- | --- | --- | --- | --- |
| GQS | 2 | 4 | 3 | 3.00 | 0.49 |
| mDISCERN | 1 | 4 | 2 | 2.20 | 0.51 |
| JAMA | 1 | 4 | 2 | 2.07 | 0.33 |
| VIQI | 5 | 11 | 7 | 7.39 | 1.15 |

**Table 4.** The evaluator used four assessment tools to evaluate the scores of the videos uploaded by non-cardiology physicians.

| Assessment Tool | Min | Max | Median | Mean | SD |
| --- | --- | --- | --- | --- | --- |
| GQS | 2 | 4 | 3 | 2.79 | 0.49 |
| mDISCERN | 2 | 4 | 2 | 2.31 | 0.54 |
| JAMA | 2 | 3 | 2 | 2.10 | 0.31 |
| VIQI | 6 | 11 | 7 | 7.21 | 1.11 |

### Video comparison with other treatment methods for coronary heart disease

To examine whether EECP-related content was comparatively underrepresented on short-video platforms, videos addressing EECP were compared with videos related to other commonly discussed treatment methods for coronary heart disease. As shown in Fig. 7, EECP-related videos accounted for a smaller proportion of CAD-related educational content compared with videos focusing on other treatment approaches.

**Figure 7.**
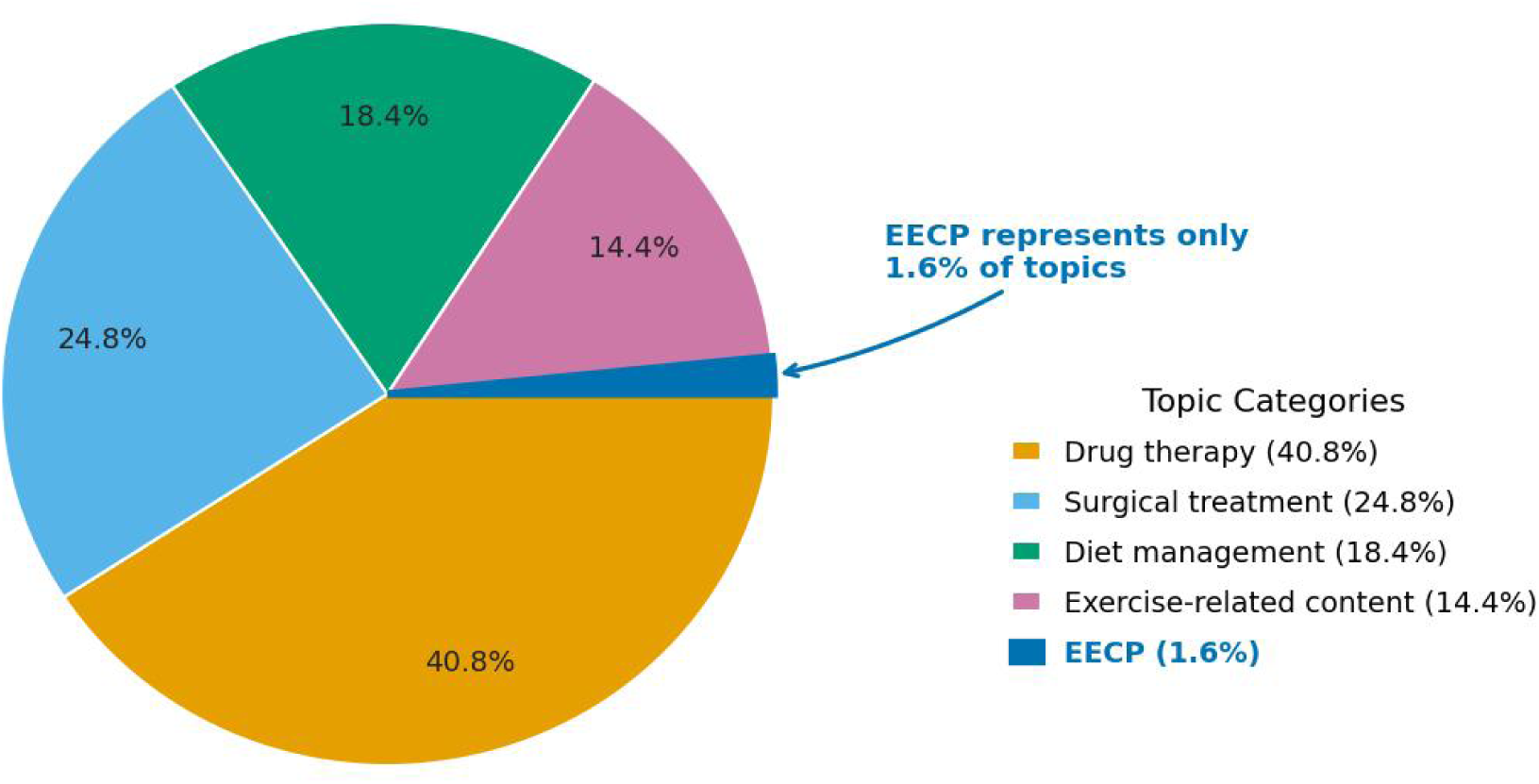
Video content distribution of CAD educational videos. (A) Likes, (B) Collections, (C) Comments, and (D) Shares. Data are presented as log10-transformed counts [log10(count + 1)]. The width of each violin represents the kernel density estimation, and the internal box plots indicate the interquartile range (IQR), with the horizontal line representing the median. Statistical significance was assessed using the Mann–Whitney U test (****P < 0.0001). (E) Radar chart comparing multidimensional informational quality between EECP-related videos and videos addressing other coronary artery disease (CAD) treatments. The axes represent four evaluation instruments: Global Quality Score (GQS), Video Information and Quality Index (VIQI), modified DISCERN (mDISCERN), and JAMA benchmark criteria. The blue polygon represents the EECP group, and the red polygon represents the other CAD treatment group. The extent of each polygon reflects relative performance across quality domains.

In terms of visibility, the number of EECP-related videos was lower than that of videos addressing alternative coronary heart disease interventions (Fig. 7). This pattern indicates that EECP-related content was less frequently represented within the short-video platform during the study period.

User engagement patterns also differed between EECP-related videos and videos focusing on other treatment methods. Comparisons of engagement metrics, including likes and comments, demonstrated that EECP-related videos generally received lower levels of user interaction than videos addressing other CAD treatments (Fig. 8). However, higher engagement was not consistently associated with higher informational quality across treatment categories.

**Figure 8.**
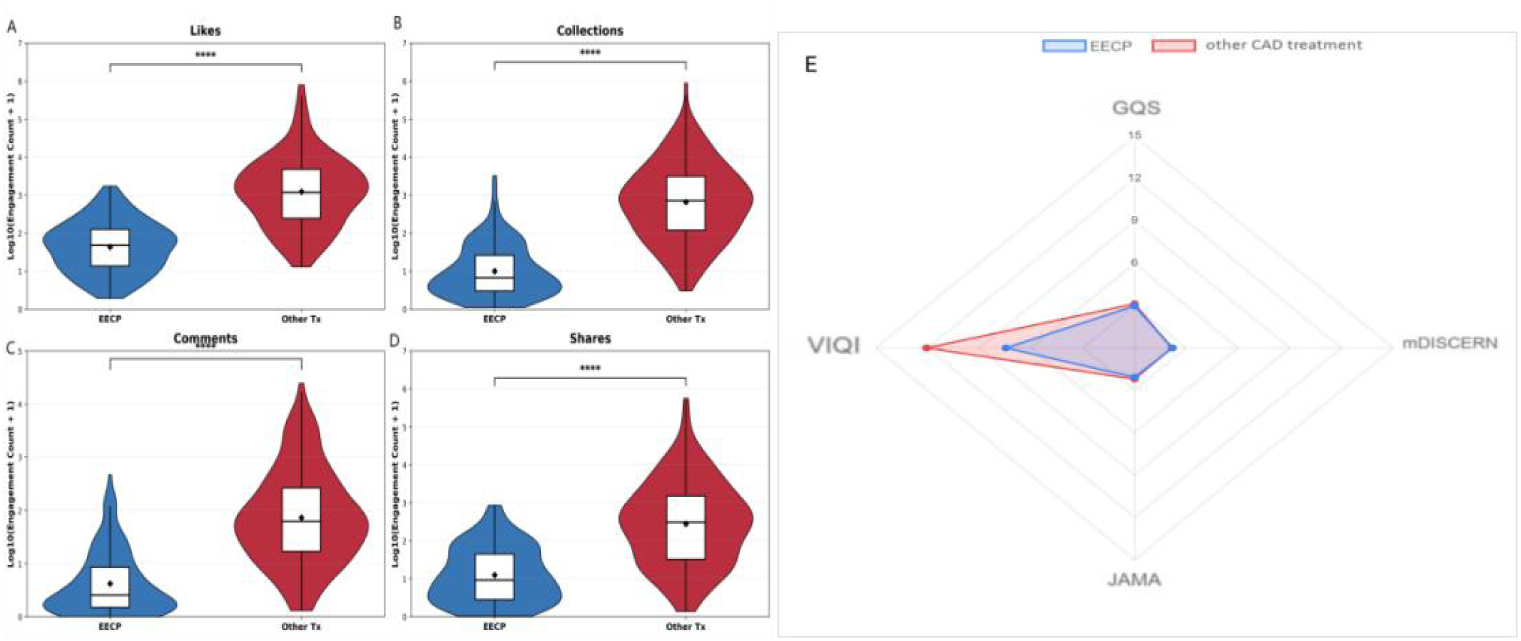
Comparison of user engagement metrics and video quality between EECP and other CAD treatment videos. The heatmaps compare the correlation patterns in “Other CAD Treatment” videos (Left Panel) versus “EECP” videos (Right Panel). Rows represent video parameters, and columns represent the four quality assessment tools.Color Coding: Red (+) indicates a statistically significant positive correlation (P < 0.05); Blue (-) indicates a statistically significant negative correlation (P < 0.05); Grey (ns) indicates no statistically significant correlation.

Correlation analyses further showed that associations between video parameters and informational quality scores varied across treatment-related video groups (Fig. 9). Overall, the patterns observed for EECP-related videos were comparable to those seen for other CAD treatment videos, with video duration showing more consistent associations with quality scores than engagement metrics.

**Figure 9.**
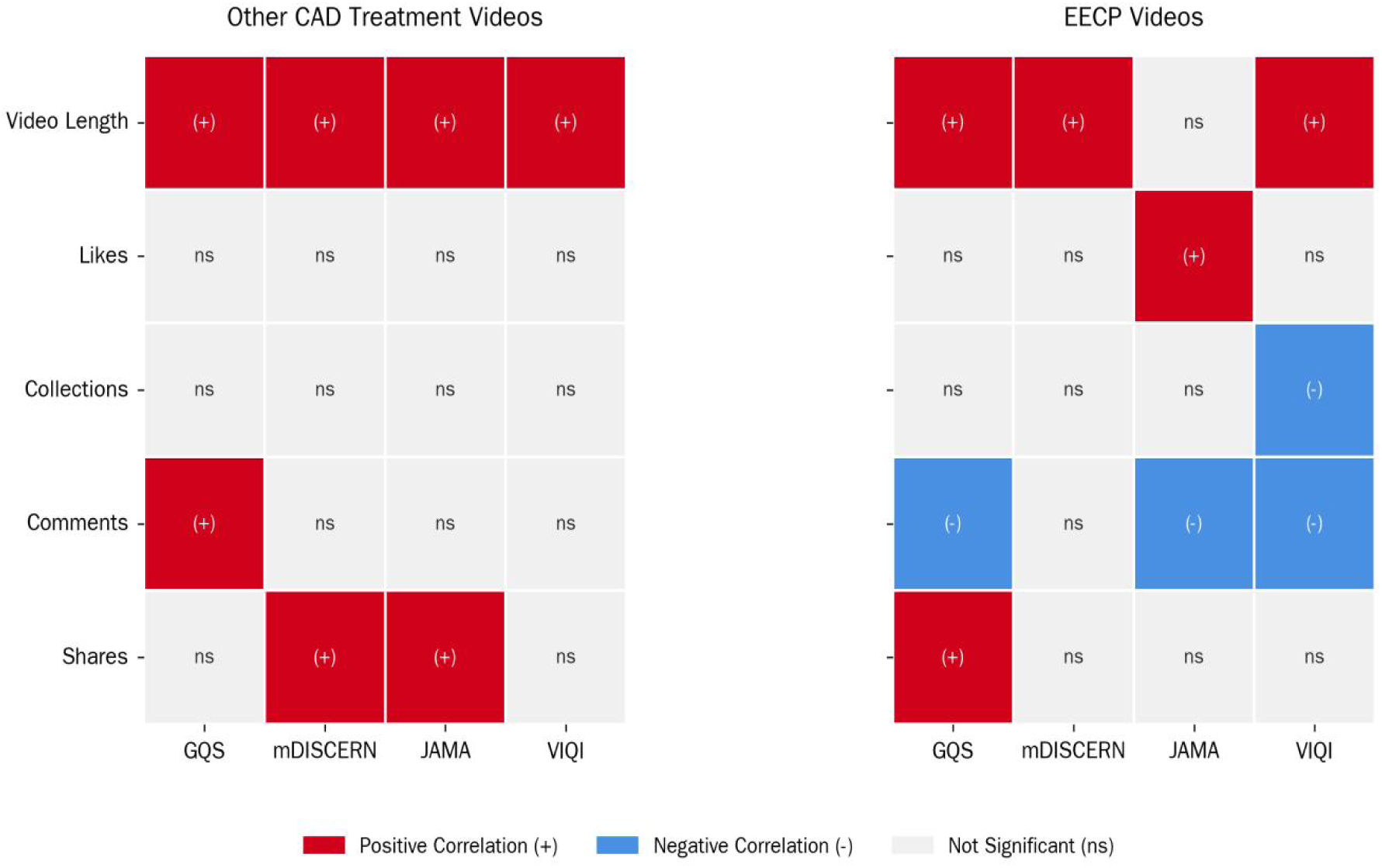
Heatmap illustrating correlations between video parameters and quality assessment scores.

Collectively, these results indicate that EECP-related videos were less prevalent and received lower user engagement compared with videos focusing on other coronary heart disease treatment methods, while the relationships between video characteristics and informational quality were broadly similar across treatment categories.

## Discussion

### Overall quality and visibility of EECP-related short videos

This study provides a comprehensive evaluation of the informational quality and dissemination characteristics of EECP–related content on a short-video platform. Using multiple validated assessment tools, we found that EECP-related videos generally exhibited low-to-moderate informational quality, were underrepresented compared with other coronary heart disease (CHD) treatment modalities, and demonstrated a notable mismatch between informational quality and user engagement. Together, these findings suggest that clinically relevant but complex therapies such as EECP may face structural challenges within short-video health communication environments [7, 32].

Across all four assessment instruments, EECP-related videos scored modestly in terms of completeness, accuracy, and overall educational value. Although some videos provided basic descriptions of EECP or shared patient experiences, detailed explanations of indications, therapeutic mechanisms, benefits, risks, and evidence-based outcomes were frequently lacking. This pattern is consistent with prior studies reporting that short-video platforms tend to favor simplified or emotionally engaging content over comprehensive medical explanations [32–34]. For therapies such as EECP, which require contextual understanding and nuanced clinical interpretation, this environment may inherently limit the depth of information conveyed.

### Mismatch between informational quality and user engagement

A key finding of this study is the weak or negative association between informational quality and user engagement metrics. Videos with higher quality scores did not consistently attract more likes or comments, whereas videos with lower informational value sometimes achieved greater engagement. This misalignment indicates that popularity on short-video platforms should not be interpreted as a proxy for informational reliability. From a public health perspective, this phenomenon raises concerns about the potential for highly visible content to convey incomplete or misleading impressions of specialized therapies [35–37].

### Video duration and informational depth

Video duration emerged as one of the most consistent predictors of higher informational quality. Longer videos were associated with improved scores across multiple assessment tools, likely reflecting greater opportunity to introduce background context, explain therapeutic mechanisms, and discuss relevant clinical considerations. However, longer duration may also reduce audience retention on platforms optimized for brief content, creating a trade-off between informational depth and viewer engagement. This tension may further contribute to the structural underrepresentation of EECP-related educational content on short-video platforms [38–39].

### Comparison with other coronary heart disease treatments

When compared with videos addressing other CHD treatments, EECP-related content was both less prevalent and less engaging. This disparity may reflect broader differences in public familiarity, patterns of clinical adoption, and perceived relevance of various treatment modalities [28]. Invasive procedures and pharmacological therapies are often more prominently featured in clinical guidelines and media narratives, whereas non-invasive adjunctive therapies such as EECP receive comparatively limited attention [40]. As a result, EECP-related content may be systematically marginalized within algorithm-driven content ecosystems.

### Implications for health communication and clinical practice

These findings have several implications for clinicians, content creators, and health communication strategists. First, reliance on short-video platforms as primary sources of information about specialized therapies should be approached with caution [41]. Second, targeted efforts by medical professionals and institutions may be necessary to improve the visibility and quality of EECP-related educational content, potentially through longer-form videos, series-based content, or integration with authoritative sources [42]. Finally, platform-level factors, including algorithmic incentives and content moderation strategies, may influence which types of medical information are most likely to reach the public [43].

### Limitations

This study has several limitations. The analysis was limited to a single platform and represents a cross-sectional snapshot in time, which may not capture temporal trends or differences across platforms. In addition, informational quality was assessed based on observable content and did not account for viewer comprehension, trust, or clinical outcomes. Despite these limitations, the use of multiple validated assessment tools and a systematic content analysis strengthens the robustness of the findings.

In conclusion, EECP-related content on short-video platforms is characterized by limited visibility, modest informational quality, and a disconnect between educational value and user engagement. These patterns suggest that complex, evidence-based therapies may be disadvantaged in short-video health communication contexts. Addressing this gap will likely require coordinated efforts from healthcare professionals, content creators, and platform stakeholders to ensure that clinically meaningful information is both accessible and accurate.

## Conclusion

This study provides a systematic evaluation of EECP-related content on a short-video platform, with a focus on informational quality, content sources, and user engagement characteristics. Overall, EECP-related videos were limited in number and demonstrated substantial variability in informational quality. Although videos produced by healthcare professionals generally achieved higher quality scores, they did not consistently receive higher levels of user engagement, indicating a persistent disconnect between informational quality and audience response in short-video environments.

Video duration was consistently associated with higher informational quality across multiple assessment tools, suggesting that structural constraints inherent to short-video formats may limit the effective communication of complex medical interventions such as EECP. In contrast, engagement metrics such as likes and comments were not reliable indicators of informational quality, underscoring the limitations of popularity-driven signals for evaluating the educational value of health-related short videos.

Compared with videos addressing other coronary heart disease treatment methods, EECP-related content appeared relatively underrepresented and less visible on the platform. This imbalance may contribute to limited public exposure to evidence-based information about EECP, despite its established clinical application.

Taken together, these findings highlight the need for targeted health communication strategies to enhance both the quality and visibility of EECP-related information on short-video platforms. Future efforts should account for platform-specific constraints and emphasize evidence-based content design to improve the dissemination of accurate and clinically meaningful information to the public.

## Data Availability

All relevant data are within the manuscript and its Supporting Information files.

